# Adolescent-Reported National Norms, Psychometrics, and Clinical Considerations for the Nationwide Quality of Life Scale (NQLS)

**DOI:** 10.64898/2026.09.25.26363826

**Authors:** Elizabeth A. Nienaber, Yinuo Liu, Eric A. Youngstrom, Mary A. Fristad

## Abstract

**Objective:** The Nationwide Quality of Life Scale (NQLS) is a brief, public-domain, youth self-report measure designed for quality of life (QoL) reporting in measurement-based care (MBC). Preliminary psychometric support comes from a sample of treatment-seeking youth, but its use in a population-based sample has not been evaluated. This study examined the NQLS’ psychometric performance in a U.S. nationally representative sample and develops national norms, score interpretation guidance, and clinical significance benchmarks.

**Method:** A nationally representative sample of 1,008 youth ages 12–18 completed the NQLS as part of a larger survey battery. Analyses included exploratory and confirmatory factor analysis, reliability, validity, item response theory, and generalized additive models for location, scale, and shape (GAMLSS) norming. We integrated these results with clinical data from 1,200 youth seeking treatment at a large Midwestern children’s hospital to estimate clinical significance benchmarks and compare between groups.

**Results:** One- and two-factor solutions were evaluated. The NQLS demonstrated good internal consistency and convergent validity, with distinguishable “life” and “school” factors. Normative percentiles were generally stable across age, with modest narrowing at the youngest and oldest ages. Normative-sample youth showed raw score and latent trait estimates significantly higher than clinical-sample youth.

**Conclusions:** The NQLS provides reliable and valid measurement of QoL in a nationally representative sample. Although two structural factors were identified, the composite score is recommended for clinical decision-making. National norms and clinical significance benchmarks may facilitate MBC applications of the NQLS. Future research should evaluate its performance across additional care settings.

---

Quality of Life (QoL) is fundamental to understanding the experiences of youth and families seeking behavioral health treatment. While the exact definition of QoL has changed over time and across disciplines, the essence remains that QoL refers to one’s satisfaction with life’s many conditions and domains (Felce & Perry, 1995; Sirgy, 2012; Wallander & Koot, 2016).

Distinct from symptom severity, monitoring QoL provides alternative perspectives to traditional concepts of progress and treatment monitoring. The Diagnostic and Statistical Manual of Mental Disorders (DSM) Strategic Committee recently announced a subcommittee focused on functioning and quality of life, which highlights the growing support for patient-centered outcomes, such as QoL, in clinical care (Oben, 2020; Brauser, 2025).

While QoL tends to be negatively correlated with symptom severity, the two constructs are not inverse; in the face of illness and stress, youth can experience levels of satisfaction comparable to healthy peers, and QoL can be low without severe symptoms (Blackwell et al., 2019). However, youth presenting with mental, emotional, and behavioral health (MEB) concerns largely report lower levels of QoL (Freeman et al., 2009; Jonsson et al., 2017; Grant et al., 2026). In behavioral health settings, QoL is a unique and important concept, as families typically present with the intention of their child “feeling better” and/or “doing better” which includes reduction of symptoms, improved satisfaction, and better functioning. QoL, “feeling better,” regaining autonomy and stability, and reducing functional impairment are examples of patient-centered outcomes, which are particularly relevant to those presenting with MEB concerns (Connell et al., 2014; Mansoor et al., 2025).

Measurement-based care (MBC) has been increasingly applied in the evaluation and treatment of MEB concerns. Across all stages of care, MBC can support accurate and thorough screening, diagnosis, and treatment or progress monitoring using patient-generated data to guide clinical decision making (Youngstrom et al., 2017). To be well-suited for MBC, a measure should be psychometrically sound, contextually appropriate, feasible, and suitable for repeated administration (*APA Professional Practice Guidelines on Measurement-Based Care*, 2025).

Reporting measure properties allows clinicians and researchers to determine its applicability throughout MBC phases, and by establishing application tools such as clinical significance benchmarks and reliable change thresholds, a measure’s utility can increase beyond screening and diagnosis to inform response to treatment, allowing within-person comparisons. As the field evolves, it is important to note that most measures used to track symptoms and support diagnosis do not also assess QoL; it is a distinct construct requiring targeted measurement that cannot be inferred from symptom severity or social characteristics (Bastiaansen et al., 2005; Jensen et al., 2022). When assessing youth with MEB concerns, QoL can be impacted across many dimensions and needs to be directly assessed (Kenny et al., 2023). Clinicians who are interested in measuring QoL should select an appropriate, dedicated, and validated measure.

The Nationwide Quality of Life Scale (NQLS) was developed to provide a brief, public-domain QoL scale for application in behavioral health. The NQLS showed initial psychometric support in a clinical sample of 828 youth receiving intensive outpatient treatment, with acceptable internal consistency, convergent validity, and sensitivity to change (Covarrubias & Fristad, 2025). Designed to fill a gap in QoL integration into MBC, the NQLS assesses life satisfaction across seven domains using seven items at a seventh grade reading level, in accordance with Hemingway’s guidance, to reduce burden on families, youth, and clinicians.

Additionally, the NQLS was designed to provide complementary information to symptom-focused measures, allowing for smooth integration into an assessment battery. Brevity, sensitivity to change, and accessibility strongly position the NQLS for uptake in behavioral health MBC (Jensen-Doss et al., 2020). However, without data on the NQLS’ performance in a population-based sample, its utility outside of a treatment-seeking sample is limited. By establishing normative scores, we can expand interpretability of NQLS scores by providing a standard for comparison across clinical settings. This allows clinicians to triage concerns, monitor outcomes, and further refine treatment plans. Additionally, with valid norms, clinicians and researchers can utilize this reference distribution to make sense of individual NQLS scores (Achenbach, 2001).

The present study aims to improve the NQLS’ utility within MBC systems by evaluating its performance in a large, non-treatment seeking, nationally representative sample of youth. Our first aim is to establish national norms for the NQLS, which will provide a frame of reference for observed scores. Our second aim is to explore the NQLS’ psychometric properties within the normative sample to ensure its factor structure, reliability, validity, and item-level discrimination and information, using item response theory (IRT) and differential item functioning (DIF) methods, outside of the original sample. We hypothesize the NQLS will show comparable factor structure to the original psychometric sample, and acceptable internal consistency and convergent validity among youth within the normative sample. Our third and final aim is to explore differences in youth-reported QoL between the normative sample and an expanded clinical sample and establish clinical significance benchmarks for direct application and increased interpretability. We hypothesize youth in the clinical sample will show lower QoL than those in the normative sample.

## Methods

### Participants

A nationally representative sample of 1,008 youth completed the NQLS as part of the larger BENCHMARK battery (osf.io/yxf8z). The sample was representative of the U.S. population on key demographics (child sex, race, ethnicity) and aligned closely with 2023 American Community Survey (ACS) estimates. Children were on average 14.4 years old (*SD* = 1.7, range = 12-18) and half (49.7%) of children identified as female. A majority (74.8%) were White, with 9.7% Black or African American, 1.9% Asian, and 14.3% Hispanic, Latino, or Spanish Origin. Nearly all parents (97.7%) had obtained a high school diploma, and 47.4% had obtained a bachelor’s degree or higher. The median household income was between $70,000 and $79,999 annually.

A subset of youth (N=90) was recontacted following their initial response to complete the battery again. The average retest interval was 19.4 days (SD=7.8). The retest sample characteristics were highly similar to those of the full sample (Supplemental Table 1), which supports the examination of test-retest dependability.

An expansion of the clinical sample used by Covarrubias and Fristad (2025) provided an additional comparator: youth seeking treatment at two intensive outpatient programs (IOPs), one focused on anxiety and one focused on mood (*N* = 1,200) in a Midwestern U.S. city. Participants ranged from 10-18 years old (14.8, *SD* = 1.5) and completed the NQLS during treatment. The clinical sample contains 372 more complete encounters than the validation sample and more data from youth historically underrepresented in behavioral health research. Although the sample is still largely white (74.7%), female (75.8%), and covered by commercial insurance (66.4%), more Hispanic, Latino, or Spanish Origin youth are present (4.6%), and the sample now reports 2.3% Asian youth. Missing data for sex, race, and ethnicity is common for electronic health record data; provided their NQLS data was complete, they were not excluded from the sample. Key demographics for the normative and clinical samples are summarized in Table 1 along with 2023 ACS estimates.

**Table 1.** Participant demographic table.

|  | Normative<br>Sample<br>(N=1008)* | Clinical<br>Sample<br>(N=1200)** | 2023<br>ACS<sup>a</sup> |
| --- | --- | --- | --- |
| **Child age** | 14.4 (1.65) | 14.8 (1.50) | - |
| 12-13 years | 34.0% | 23.8% | - |
| 14-15 years | 35.2% | 40.8% | - |
| 16-18 years | 30.8% | 34.8% | - |
| **Child sex** |  |  |  |
| Female | 49.7% | 75.8% | 50.5% |
| Male | 50.3% | 23.6% | 49.5% |
| **Race** |  |  |  |
| White | 74.8% | 74.7% | 60.5% |
| Black or African American | 9.7% | 10.0% | 12.1% |
| Asian | 1.9% | 2.3% | 6.0% |
| American Indian or Alaska Native | 0.8% | 0.0% | 1.0% |
| Native Hawaiian or Pacific Islander | 0.2% | 0.0% | 0.2% |
| Some other race | 3.3% | 0.6% | 7.4% |
| Multiple races selected | 8.9% | 8.6% | 12.8% |
| Missing | - | 3.8% | - |
| **Ethnicity** |  |  |  |
| Hispanic, Latino, or Spanish Origin | 14.3% | 4.6% | 19.4% |
| Not Hispanic, Latino, or Spanish Origin | 85.7% | 85.8% | 80.6% |
| Missing | - | 9.6% | - |
| **Parent/guardian education** |  |  |  |
| High school graduate or higher | 97.7% | - | 89.8% |
| Bachelor's degree or higher | 48.7% | - | 36.2% |
| **Insurance Status** |  |  |  |
| Commercial | 64.1% | 66.4% | 67.0% |
| Medicaid | 35.3% | 28.5% | 37.4% |
| Other | 9.4% | 3.0% | 5.4% |
| Missing | - | 2.0% | - |
| **Median Household Income** | \$70,000-\$79,999 | - | \$77,719 |

<sup>a</sup>Data from 2023 American Community Survey (ACS) 1-year estimates.
\*2 normative-sample youth reported intersex and were excluded from sex-based analyses
\*\*0.6% of the clinical sample youth were aged 10-11

### Procedure

The BENCHMARK study was developed and executed by researchers at a pediatric academic health center in a Midwestern U.S. city who collected data on a sample of youth-parent dyads to better understand the prevalence of MEB symptoms across the country. Data collection was conducted by YouGov, an external survey agency with a large, global panel of members and the capability to conduct nationally representative surveys. YouGov first polled parents of 12– 18-year-olds, then their respective child, to complete the BENCHMARK assessments. Initial data cleaning and stratification was performed by YouGov to align the sample with the 2023 ACS estimates on sex, race, ethnicity, parent education level, and household income. Final cleaning and all analyses were conducted by the authorship team. This study was approved by the local Institutional Review Board (IRB) and granted a waiver of consent as all participant data received by the hospital were minimally identifiable.

The clinical sample is a secondary data set exempt by the institution’s IRB as collection of NQLS scores are related to hospital quality improvement and program evaluation. The NQLS was administered at IOP intake and discharge. Only initial NQLS scores were considered in this study; youth with incomplete data were listwise deleted. Data were collected from the electronic health record and cleaned by the authorship team.

### Measures

#### Quality of Life

The Nationwide Quality of Life Scale (NQLS) is a seven-item scale that assesses satisfaction among seven life domains: friends, family, grades, school in general, health, life, and anticipated future. Originally developed for youth self-report (Covarrubias & Fristad, 2025), the NQLS has since been evaluated for parent-report within a large nationally representative community sample (P-NQLS; Liu et al., 2026). Each item is scored on a five-point Likert scale (0 = very unsatisfied to 4 = very satisfied); the total scores range 0-28, where higher scores indicate higher youth QoL (NQLS) or parent perception of higher youth QoL (P-NQLS). ***Symptom Severity***

#### Concise Health Risk Tracking Scale (CHRT)

A psychometrically reliable and valid measure of suicide risk and associated factors, research on the CHRT supports its use in adolescents experiencing suicidality. This study utilizes the 16-item CHRT, which builds on the traditional six-item suicidal thoughts and propensity subscales to include two items assessing impulsivity and two items assessing irritability (Trombello et al., 2023), both of which are associated with adolescent mood impairment (Leibenluft et al., 2024; Van Meter et al., 2016).

### Very Quick Inventory of Depressive Symptomatology (VQIDS)

A five-item measure of core depression symptoms, the youth self-report VQIDS is adapted from the previous interview-style Quick Inventory of Depression Symptomatology (QIDS; Trivedi et al., 2004). The VQIDS shows reliable and valid measurement of depressive symptoms in youth (De La Garza et al., 2017).

#### Pediatric Symptom Checklist-17 (PSC-17)

The PSC-17 measures internalizing, externalizing, and attention symptoms along three subscales (Gardner et al., 1999). Commonly used in screening contexts to identify youth who would benefit from additional evaluation, the measure is well suited for MBC applications due to its brevity and specificity.

#### Vanderbilt ADHD Diagnostic Rating Scale (VADRS)

The VADRS is an ADHD rating scale which covers all DSM symptoms of ADHD across two nine-item subscales covering hyperactivity-impulsivity and inattention (Wolraich et al., 2003). Typically used as a parent or teacher-report measure, we examine self-reported ADHD symptoms to evaluate their relationship with QoL without introducing rater discrepancies.

### Impairment and Functioning

**Nationwide Gauge of Emotional Severity (N-Gauge).** A single-item scale, currently in development, which assesses youth global emotional functioning on a scale from 0-5 (McClellan et al., 2025; Supplemental Figure 1).

**Vanderbilt ADHD Diagnostic Rating Scale (VADRS).** In addition to the symptom-severity subscales, the VADRS also directly assesses impairment using a seven-item performance section. This asks youth to report on how their symptoms are directly impacting their functioning.

### Statistical Analysis

All analyses were conducted in RStudio using the *tidyverse, lavaan,* and *mirt* packages.

All visualizations were created in RStudio using the *ggplot2* and *semPlot* packages.

### National Norms

We established age- and sex-specific continuous norms for the NQLS with Generalized Additive Models for Location, Scale, and Shape (GAMLSS; (Rigby et al., 2017). Guided by Timmerman et al. (2021), we compared multiple candidate distributions and selected the best-fitting model based on the lowest GAIC and BIC values as well as global deviance and overall parsimony (Supplemental Table 2). The final model used a Box-Cox Cole and Green (BCCG) distribution with P-spline smoothers to capture age-by-sex interactions in mean (μ) and age-dependent changes in variance (σ), while holding skewness (ν) and kurtosis (τ) constant. The worm plot (Buuren & Fredriks, 2001) confirmed adequate local fit (Supplemental Figure 2). Based on the GAMLSS output, we estimated smoothed percentile curves and tables (5^th^, 10^th^, 25^th^, 50^th^, 75^th^, 90^th^, 95^th^) separately for males and females.

### Psychometrics

**Factor Analysis.** We first performed exploratory factor analysis (EFA) using polychoric correlations and weighted least squares with promax rotation. Polychoric correlations were selected due to the ordered, Likert-type response format of the NQLS (Olsson, 1979). Factor retention criteria considered include eigenvalues and optimal coordinates (Raîche et al., 2013), scree plot, minimum average partial (MAP), and parallel analysis using principal components extraction (Horn, 1965; Auerswald & Moshagen, 2019). We then performed confirmatory factor analyses (CFA) and exploratory structural equation modeling (ESEM) using the recommended weighted least squares mean and variance (WLSMV) estimators and standard geomin rotation to further delineate factor loadings and cross-loadings (Li, 2016; Muthén et al., 1997). Model structural fit was evaluated using the comparative fit index (CFI), Tucker-Lewis index (TLI), root mean square error of approximation (RMSEA, with 90% confidence interval), standardized root mean square residual (SRMR), and a factor loading threshold of .30. Univocality was explored using first-second eigenvalue ratios, explained common variance, and item correlations. Candidate models were refitted using robust maximum likelihood (ML) estimation, treating responses as approximately continuous, to obtain Akaike Information Criterion (AIC) and Bayesian Information Criterion (BIC) values for supplementary model comparison.

**Internal Consistency.** Internal consistency was evaluated using both Cronbach’s alpha (α; Cronbach, 1951) and McDonald’s omega (ω; McDonald, 1999). Due to the length of the NQLS, we hypothesize alpha will underestimate true reliability while inflating reliability attributable to a *g*-factor due to the assumption of tau-equivalency (Revelle & Condon, 2019); both α and total ω are reported, but emphasis is placed on ω. Reporting inter-item correlation further supports internal consistency without sample-size bias (Streiner et al., 2015).

**Test-Retest Dependability.** Test-retest dependability of NQLS total scores between the original and retest administrations was examined using intraclass correlation coefficient (ICC) and Bland-Altman plots. ICC, specified as a two-way random effects model with absolute agreement and single unit of analysis (ICC[A,1]), estimates absolute score stability over time. The Bland-Altman plot (Bland & Altman, 1986) depicts the difference between the test and retest NQLS scores (y-axis) against their mean scores (x-axis), visualizing the agreement between the two administrations. We also conducted a Johnson-Neyman analysis to identify a region of statistically distinguishable difference.

**Convergent Validity.** Pearson’s correlations between the NQLS and other symptom severity, impairment, and functioning measures were calculated to examine the NQLS’ convergent validity, with Benjamini-Hochberg corrections applied.

**Item Response Theory.** We estimated a correlated two-dimensional graded response model (GRM) and separate unidimensional GRMs for factor-specific parameters and conditional precision. In addition to the correlated and univocal models, a unidimensional GRM containing all seven items was estimated to analyze the composite NQLS score (Samejima, 1969). All models include standard error for estimated parameters and were evaluated for fit using the *C_2_* and *M_2_*statistics, CFI, TLI, RMSEA, and SRMSR. The correlated and composite models, both containing all seven NQLS items, were compared using AIC and BIC values. Local independence was examined using Yen’s Q3 residual correlations (Yen, 1984). Item fit was assessed using binned RMSD diagnostics. Items with RMSD values above .10 were flagged for potential misfit (Yamamoto et al., 2013). Person fit was evaluated using the standardized *Z_h_* statistic (Drasgow et al., 1985) with | *Z_h_* | > 3 indicating an atypical response pattern. Conditional precision was evaluated test-level using information, standard error, and conditional reliability curves. Item information curves are presented.

**Differential Item Functioning.** To explore the generalizability of these findings, we then used the correlated GRM estimated above to check for differential item functioning (DIF) by sex (male vs. female), age (12-14 vs. 15-18), and race (White vs. non-White) within the normative sample using hybrid ordinal logistic regression and IRT in *lordif*. Items were flagged for DIF if McFadden’s *R^2^* showed a change greater than .02 between models, a standard effect size threshold for DIF detection. DIF was not evaluated for the two-item ’school’ factor across any strata, as it does not meet *lordif*’s minimum item requirement.

### Clinical Sample Evaluation and Application

Using the expanded clinical dataset, we performed CFA as a replication of the previous clinical findings by Covarrubias and Fristad (2025). Following the guidance of Millsap and Tein (2004), we evaluated measurement invariance at the configural, metric, and scalar levels to determine if claims could be made about latent mean differences between clinical and normative sample youth (Putnick & Bornstein, 2016). Independent sample t-tests were utilized to identify significant group differences (clinical vs. normative youth) in item endorsement rates or latent trait differences with Benjamini-Hochberg corrections applied. Effect size for latent trait differences is reported in normative *SD* units.

Clinical significance benchmarks were established to determine when a youth’s self-reported QoL may be showing clinically significant improvements or deteriorations based on the reliable change index, calculated using the standard error of the difference for raw scores reported by normative-sample youth (Jacobson & Truax, 1991). Additionally, we calculated the minimally important difference (MID), which identifies the lowest possible score change that is associated with personally or clinically noticeable change in symptoms, which may or may not be statistically meaningful, approximated as half a standard deviation of the scale (Guyatt et al., 1993; Streiner et al., 2015). Ninety and 95% reliable change (RC) scores were calculated to provide statistically significant indicators of change, more sensitive than the MID. We used the NQLS’ 2-week test-retest dependability to calculate RC scores to examine genuine fluctuation in reported QoL across occasions in a way that an internal consistency estimate cannot (Revelle & Condon, 2019). Together, the MID, RC scores, and clinical significance benchmarks provide clinical applications and interpretations for NQLS scores.

## Results

### BENCHMARK Sample

#### National Norms

The range of NQLS scores showed a slight narrowing as age increases across sex, with the 5^th^ percentile reporting higher QoL, and the 95^th^ percentile reporting lower QoL as age increases. Females in the 5^th^ percentile reported a total score of 12 at 12 years old and a total score of 13 at 18 years old. Males in the 5^th^ percentile reported a total score of 12 across all ages. Females in the 95^th^ percentile reported a total score of 27 at 12 years old and a total score of 25 at 18 years old. Males in the 95^th^ percentile reported a total score of 26 at 12 years old and a total score of 25 at 18 years old. Percentile curves provide a visual demonstration of this narrowing and depict the consistency of the 50^th^ percentile over time (Figure 1); there was no observed change in NQLS total score as age increased across average males (50^th^ percentile = 20) or females (50^th^ percentile = 21). Percentile tables show slight differences between male- and female-reported QoL with females showing equal or higher NQLS total scores across all ages (Table 2). Overall, NQLS total scores remained relatively stable across ages within each percentile, showing minimal age-related change in what could be interpreted as ‘typical’ for youth QoL.

**Figure 1.**
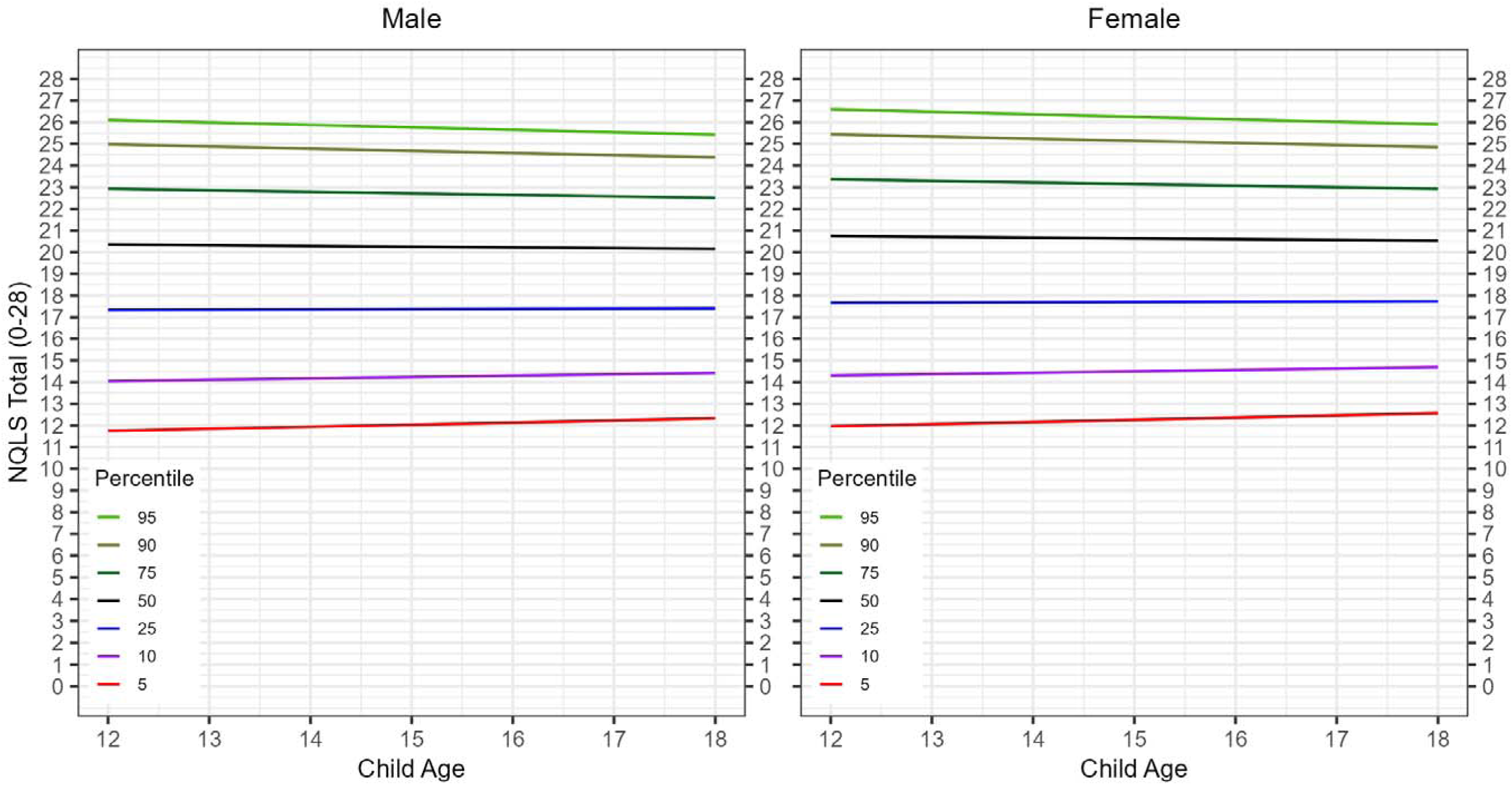
GAMLSS-estimated sex-stratified percentile curves (5^th^-95^th^) for Nationwide Quality of Life Scale (NQLS) self-report total scores (range 0-28; ages 12-18).

**Table 2.**
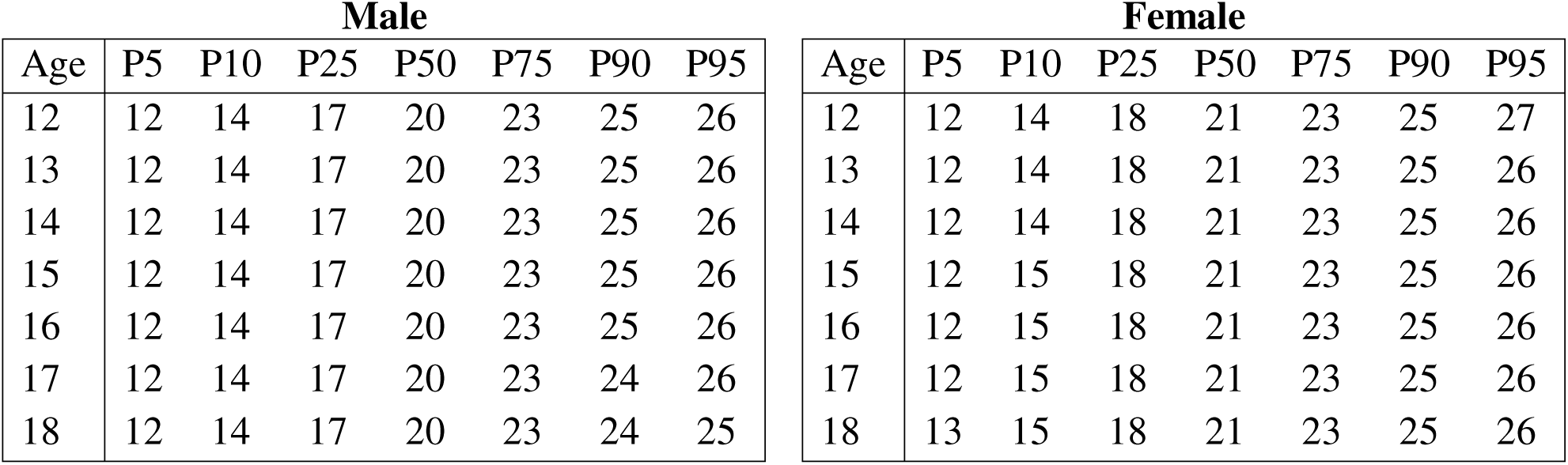
Sex-stratified normative percentiles of Nationwide Quality of Life Scale (NQLS) total scores reported by 12-18-year-olds.

| Male |  |  |  |  |  |  |  | Female |  |  |  |  |  |  |  |
| --- | --- | --- | --- | --- | --- | --- | --- | --- | --- | --- | --- | --- | --- | --- | --- |
| Age | P5 | P10 | P25 | P50 | P75 | P90 | P95 | Age | P5 | P10 | P25 | P50 | P75 | P90 | P95 |
| 12 | 12 | 14 | 17 | 20 | 23 | 25 | 26 | 12 | 12 | 14 | 18 | 21 | 23 | 25 | 27 |
| 13 | 12 | 14 | 17 | 20 | 23 | 25 | 26 | 13 | 12 | 14 | 18 | 21 | 23 | 25 | 26 |
| 14 | 12 | 14 | 17 | 20 | 23 | 25 | 26 | 14 | 12 | 14 | 18 | 21 | 23 | 25 | 26 |
| 15 | 12 | 14 | 17 | 20 | 23 | 25 | 26 | 15 | 12 | 15 | 18 | 21 | 23 | 25 | 26 |
| 16 | 12 | 14 | 17 | 20 | 23 | 25 | 26 | 16 | 12 | 15 | 18 | 21 | 23 | 25 | 26 |
| 17 | 12 | 14 | 17 | 20 | 23 | 24 | 26 | 17 | 12 | 15 | 18 | 21 | 23 | 25 | 26 |
| 18 | 12 | 14 | 17 | 20 | 23 | 24 | 25 | 18 | 13 | 15 | 18 | 21 | 23 | 25 | 26 |

#### Psychometrics

**Factor Structure.** Before proceeding with psychometric analyses, we evaluated the data’s suitability for factoring and psychometric analysis. The Kaiser-Meyer-Olkin test found adequate sampling and variance with an overall measure of sampling adequacy (MSA) of .83 and item-level MSA ranging from .78 to .91. Bartlett’s test of sphericity showed statistical significance, χ*²* (21) = 2957.64, *p* <.001, indicating sufficient variable correlations to proceed.

Initial factor retention criteria favored a one-factor solution. The scree plot showed a significant elbow after retaining the first factor, with parallel analysis (principal-components extraction), optimal coordinates, and the minimum average partial criterion supporting the retention of one factor. EFA global fit indices suggested a two-factor solution, with the one-factor EFA showing adequate fit, χ*²* (14) = 238.07, TLI = .80, RMSR = .07, RMSEA = .17, but by forcing a two-factor EFA, global fit improved across all indices, χ*²* (13) = 33.23, TLI = .93, RMSR = .03, RMSEA = .10. In the one-factor solution, items showed adequate loading onto the single factor, ranging from .56 (grades) to .88 (life). In the two-factor solution, items 3 and 4 break away to form the second factor, “school,” with loadings of .82 for satisfaction with grades and .76 for satisfaction with school, while the other five items loaded onto a broader “life” factor. In the two-factor solution, the loading of item 6 (life) increased to 1.02; Heywood’s warning indicates that the two-factor solution should be interpreted cautiously.

One-factor and two-factor CFA were estimated to further compare the competing fit indicated by EFA. The one-factor composite model demonstrated mixed fit, χ*²* (14) = 414.26, CFI = .92, TLI = .88, RMSEA = .17, SRMR = .07, with substantially stronger fit indicated for the two-factor correlated model, χ*²* (13) = 67.47, CFI = .99, TLI = .98, RMSEA = .06, SRMR = .03 (Supplemental Table 3). Satisfaction with school and grades load strongly onto the “school” factor (range = .74-.86), while the other 5 items load moderate-to-strongly onto the “life” factor (range = .59-.93). The “school” and “life” factors correlate .67, indicating a strong relationship, but distinct constructs. Individual factor loadings for each solution are presented in Supplemental Table 4.

The bifactor CFA did not converge, most likely reflecting fragility of the two-item “school” factor due to limited item identification. As a fallback, a Schmid-Leiman decomposition (psych::omega()) estimated omega-hierarchical (ωH) and explained common variance (ECV); because only two group factors were specified, this required constraining the general factor’s loadings on both group factors to be equal for identification (a standard workaround, not a free estimate), and the resulting values (ωH = .66, ECV = .64, ωtotal = .88) should be interpreted with that caveat as opposed to a fully identified estimate. Average relative parameter bias and percentage of uncontaminated correlations still require a proper bifactor decomposition and remain inadmissible under this workaround. Remaining descriptive indices provided evidence of substantial unidimensionality across a broad ‘QoL’ dimension: for the total item set, the first-second eigenvalue ratio (EV1/EV2) was 3.93, with the first factor explaining 53.3% of the variance, and corrected item-total correlations ranging from .47 to .69. For the “life” items EV1/EV2 was 4.4, with 60.5% of variance explained by the first factor, and corrected item-total correlations ranging from .48 to .72; “school” items showed an EV1/EV2 = 4.47, with 81.7% of variance explained by the first factor, and the inter-item correlation = .64. As the EV1/EV2 and variance-explained statistics are reflective of a single interitem correlation, they should not be interpreted as evidence that the “school” factor is a sufficiently established subscale. Collectively, these findings supported the presence of a broad QoL dimension but did not establish essential unidimensionality for the full seven-item set, nor sufficient psychometric support for independent subscales.

A two-factor ESEM was estimated to determine if allowing cross-loadings would alter model stability and meaningfully change the structural interpretation. With marginally better global fit compared to the two-factor correlated CFA, χ*²* (8) = 25.19, CFI = .99, TLI = .99, RMSEA = .05, and SRMR = .02, the model was considered overfit to our data, evidenced by limited benefit from added complexity (Supplemental Table 3). Supplementary maximum-likelihood estimations produced slightly lower AIC in the ESEM compared to the two-factor CFA (16,289.71 versus 16,295.97), but BIC values favored the parsimonious two-factor CFA (16,404.11 versus 16,422.44). ESEM cross-loadings are presented in Supplemental Figure 3. As the improvement in global fit did not change the loading pattern or interpretation of the two factors, the two-factor correlated CFA was retained as the preferred model for the NQLS’ internal structure.

Interpretation of these findings requires the differentiation of the NQLS’ measurement structure from the recommended clinical scoring approach. Fit indices showed that a two-factor structure with correlated “life” and “school” factors is the best representation of the internal structure. However, with only two items strongly loading onto the “school” factor, our ability to perform subscale-specific psychometric diagnostics was limited. The evidence available to interpret the “school” factor as a standalone score did not support its precision or coverage of school-related satisfaction, making clinical interpretations potentially misleading. In combination with the strong factor-level correlations between “life” and “school” and presence of a broad QoL dimension, this limitation supported the seven-item total score’s utility as a clinically useful composite. Thus, the total score is recommended for individual clinical interpretations and used as the basis for clinical application analyses, including norms and clinical significance benchmarks. The two-factor correlated structure is used for research or group-level analyses.

This recommendation is based on clinical interpretability and reliability, validity, and precision of the total score, although the two-factor model fits the data better globally. Both are used throughout this paper, but analyses will be clearly indicated as based on either the composite model (all 7 items loading onto the “QoL” factor) or the correlated model (items 3 and 4 loading onto the “school” factor; items 1, 2, 5, 6, and 7 loading onto the “life” factor).

**Internal Consistency.** The NQLS total score demonstrated good internal consistency (α = .81, correlated model-based ωtotal = .88). The five-item “life” factor shows internal consistency similar to the total score, α = .78; ωtotal = .84. Cronbach’s alpha and Spearman-Brown reliability estimates of the two-item “school” factor should be interpreted with caution due to the low number of indicators on the factor (α = .73; *r_s_* = .78). Inter-item correlation of .38 supports the presence of distinct but related items.

**Test-retest Dependability.** Intraclass correlation, raw score difference, and Bland-Altman analyses further characterized agreement between baseline and retest administrations of the total score (N = 90, strict-matched) – consistent with this study’s recommendation that the total score, not the two-factor structure, be used for individual-level clinical interpretation (see Factor Structure, above). Mean scores were 19.51 (*SD* = 4.39) at baseline and 20.29 (*SD* = 4.18) at retest. Absolute agreement was ICC(A,1) = .72 [95% CI: .60, .81]. Scores increased slightly from baseline to retest (*Mdiff* = −.78 points, *d_z* = −.25, *t*(89) = −2.33, *p* = .022), the same regression-to-the-mean or practice-related pattern observed in this initiative’s companion suicide-risk measures, appearing here as a small apparent improvement because the NQLS is scored in the opposite direction (higher scores reflecting better functioning).

Bland-Altman analysis found no evidence of proportional bias (*b* = .06, *p* = .50), indicating the magnitude of the baseline-retest difference did not vary systematically with overall QoL. A Johnson-Neyman analysis identified a region of statistically distinguishable difference from zero within the middle of the observed range (14.9-21.3), consistent with greater estimate precision where the sample is densest rather than a severity-dependent effect (Figure 2).

**Figure 2.**
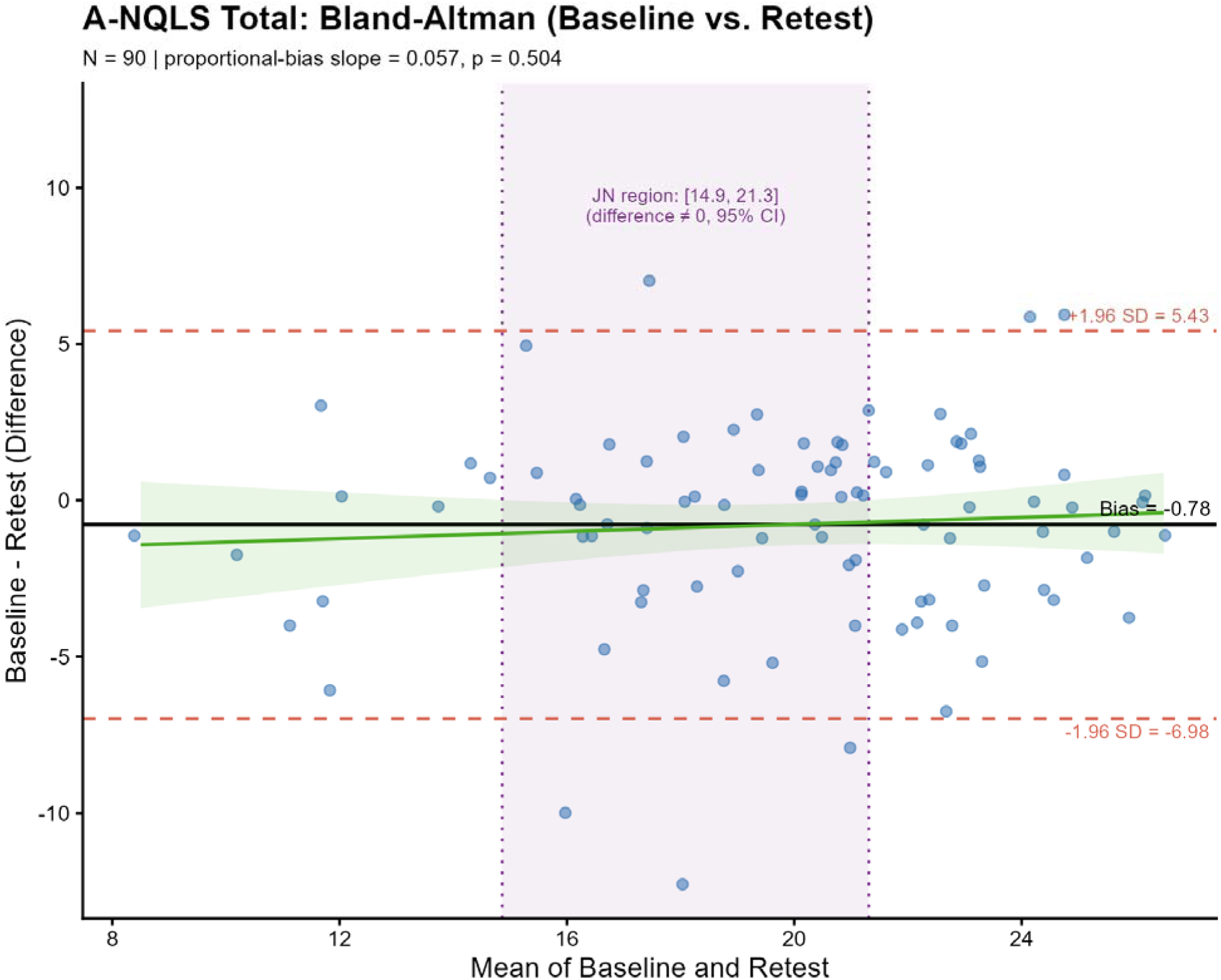
Patterns of retest agreement visualized as a Bland-Altman plot with region of significance (N = 90).

**Convergent Validity.** Correlations with measures of impairment, symptom severity, and global emotional functioning provide evidence for convergent validity of the NQLS within a nationally representative sample. All measures used in this study were youth self-report, and all total score correlations were negative and significant beyond *p* < .001 with 95% confidence intervals provided for each relationship (Table 3).

**Table 3.**
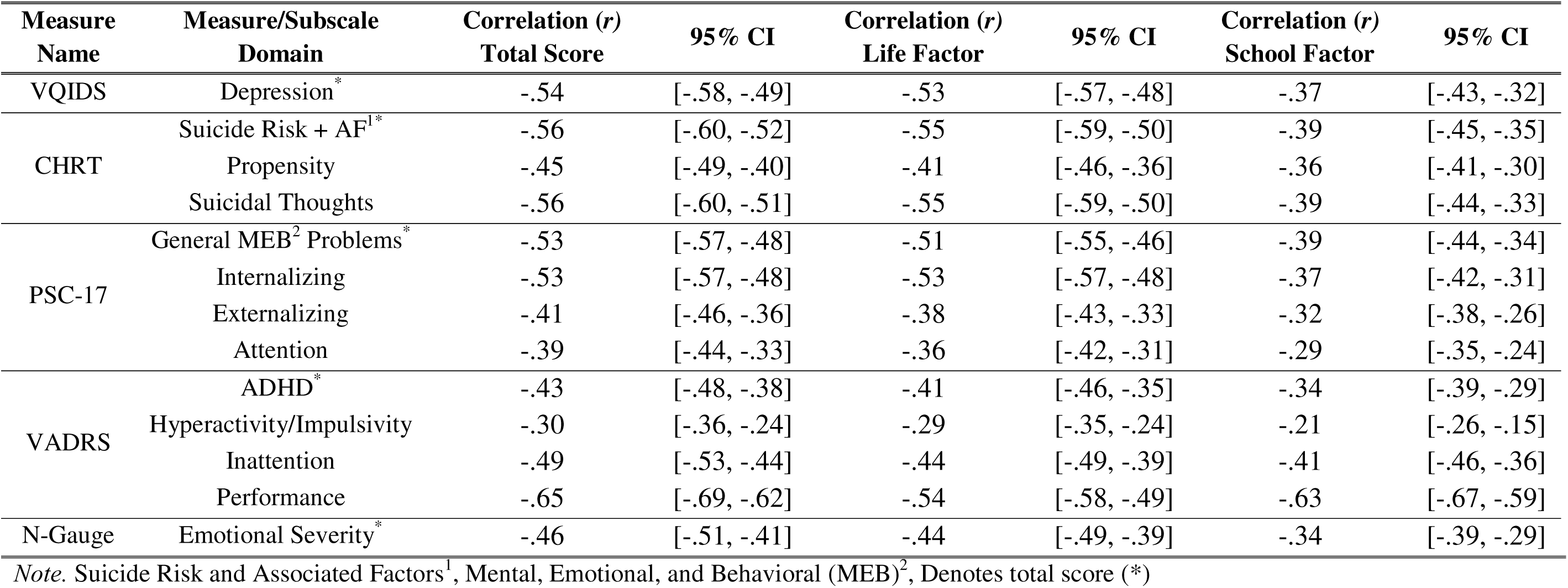
Correlation between Nationwide Quality of Life Scale (NQLS) and other BENCHMARK measures.

As expected, higher QoL as measured by NQLS total score was associated with lower impairment and symptom severity. The strongest relationship was observed between NQLS total score and VADRS-performance score (*r* = −.65). Moderate negative correlation was observed between NQLS total score and total scores of the VQIDS (*r* = −.54), CHRT (*r* = −.56), and PSC-17 (*r* = −.53), as well as the CHRT’s suicidal thoughts subscale (*r* = −.56) and the internalizing subscale of the PSC-17 (*r* = −.53). Weak but significant correlations were observed between NQLS total score and the propensity subscale of the CHRT (*r* = −.45), the externalizing and attention subscales of the PSC-17 (*r* = −.41; −.39), and all symptom scales of the VADRS (total score, *r* = −.44; inattention, *r* = −.49; hyperactivity-impulsivity, *r* = −.30). A significant correlation was found between the N-Gauge and NQLS total score (*r* = −.46); this measure is being evaluated for exploratory purpose only. Overall, this pattern indicates the NQLS total score was most strongly related to functional impairment and internalizing symptoms, with weaker associations to externalizing and hyperactivity symptoms.

All factor-level correlations were negative and significant beyond *p* = .001, with the “life” factor most strongly associated with suicidal thoughts and suicidal risk and associated factors measured by the CHRT (*r* = −.55), and the VADRS performance items most strongly correlated with the “school” factor (*r* = −.63). The “life” factor correlates with symptom severity measures more consistently; correlations ranged from −.29 (VADRS hyperactivity/impulsivity subscale) to −.55 (CHRT suicide risk and suicidal thoughts subscales). The “school” factor showed more variation, with correlations ranging from −.21 (VADRS hyperactivity/impulsivity subscale) to −.63 (VADRS performance subscale).

We evaluated differences in dependent correlations using Steiger’s test (*Z*), with Cohen’s *q* quantifying the magnitude of the difference. False-discovery-rate correction was applied using the Benjamini-Hochberg procedure. Correlations between the NQLS total score and “life” score were comparable, with few producing meaningful effect sizes. However, when compared to the “school” score, both total score and “life” score demonstrated stronger negative associations with internalizing and suicide-related constructs. For example, the “life” factor was more strongly correlated with depression as measured by the VQIDS than the “school” factor, *q* = −.21, *Z* = − 9.12, *p*_FDR_ < .001. The “school” factor demonstrated stronger association with performance impairment compared to the “life” factor using the VADRS performance total score (*q* = .14, *Z* = 3.78, *p*_FDR_ < .001). Several statistically significant differences were found, but Cohen’s *q* indicated no more than small effect sizes (Supplemental Table 5).

**Item Response Theory.** Within the normative sample, an inspection of discrimination parameters showed consistency with the two-factor internal structure. Items 1, 2, 5, 6, and 7 had their primary discrimination parameter on the “life” factor, where items 3 and 4 had their primary discrimination parameters on the “school” factor. The latent dimensions were strongly correlated (*r* = .73). Between the two GRMs containing all seven items, the two-factor correlated model produced better relative fit compared to the composite model, evidenced by lower AIC (38365.71 vs. 38766.88), BIC (38570.92 vs. 38966.39), and sample-size adjusted BIC values (38456.54 vs. 38855.19; Supplemental Table 6). The composite and correlated models cannot be directly compared to the “school” and “life” factor-level GRMs as they were estimated using different item sets. Primary discrimination parameters ranged from .92 to 4.32 for the “life” factor, and from 2.21 to 3.38 for the “school” factor. Category thresholds were ordered as expected across all items, indicating progressively higher response categories correspond to higher levels of the latent trait. Due to the limited number of indicators on the “school” factor, its precision should be interpreted with caution.

At the item level, the mean Q3 residual correlation was −.13 for the two-factor correlated model, ranging from −.53 to .12. Friends and family showed notably elevated positive residual dependence (Q3 = .12), which indicates possible local dependence beyond that of the latent dimensions of “life” and “school.” Correlated binned RMSD values ranged from .05 to .11 with only “school” exceeding the .10 threshold. Detailed item-level fit results are presented in Supplemental Table 7. At the person level, 19 respondents had absolute person-fit values above threshold, |*Z_h_*| > 3.

The “life” dimension provided its greatest measurement information between approximately θ = −2.15 and θ = 1.80, with conditional reliability exceeding .80 from θ = −2.87 to θ = 1.39. “School” provided its greatest information between θ = −1.60 and θ = 2.10, with conditional reliability exceeding .80 from θ = −2.41 to θ = 1.10. In the unidimensional composite model provided its greatest information between approximately θ = −2.40 and θ = 2.05, with conditional reliability exceeding .80 across θ = −3.42 to θ = 1.57 (Figure 3). Differential item information functions are presented in Figure 4. Though the correlated two-factor model showed more favorable structural evidence, the composite GRM provided complementary information regarding the precision of the seven-item total score. The total score showed reliable measurement across a wide range of the latent trait, though precision declined among those reporting high QoL.

**Figure 3.**
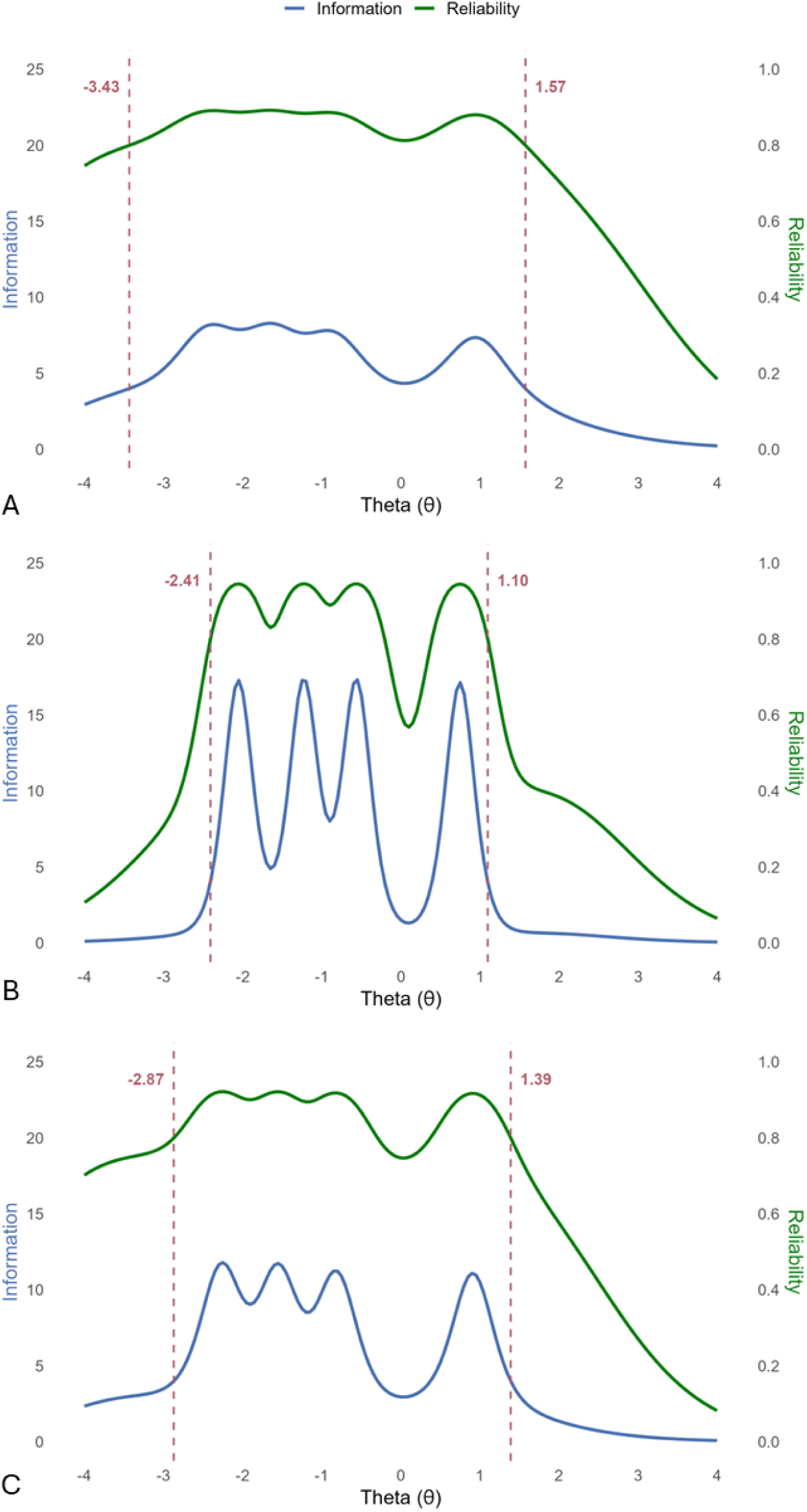
Conditional reliability of Nationwide Quality of Life Scale (NQLS) composite score (A), “school” factor (B), and “life” factor (C).

**Figure 4.**
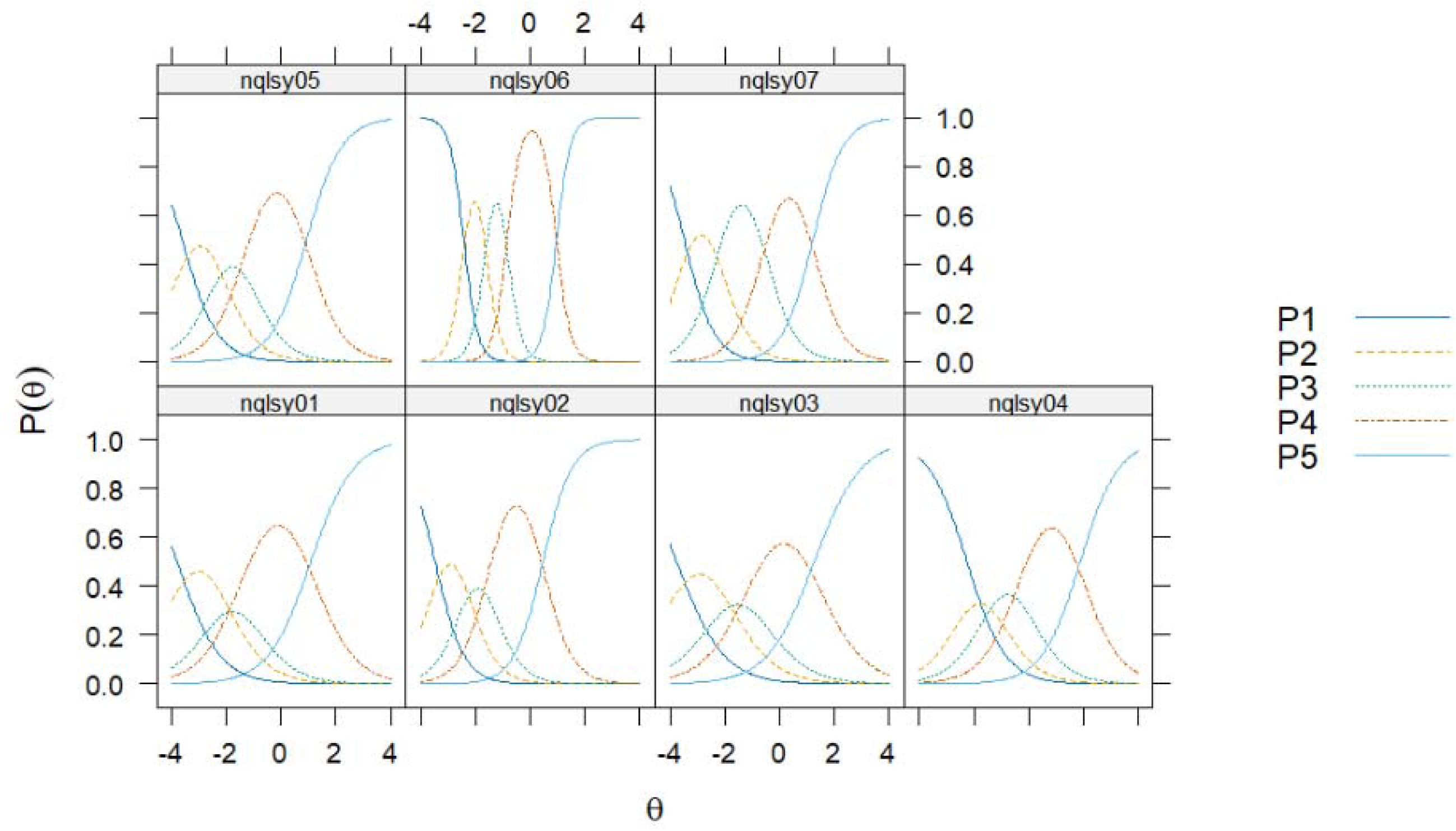
Nationwide Quality of Life Scale (NQLS) composite graded response model item characteristic curves.

**Differential Item Functioning.** Item-level DIF was evaluated for the total score and the “life” subscore using the same iterative hybrid ordinal logistic regression procedure (*R²* change ≥ .02, α = .01). No items were flagged for significant DIF across race, sex, or age strata, with all pseudo-*R²* change values below .005 except one; item 6 showed slightly elevated *R²* for White vs. non-White youth in comparison to the other items but was still well below the identified threshold (*R²* = .007).

## Clinical Sample Replication and Application

### Psychometric Extension

This study observed adequate sampling (MSA = .77) and internal consistency (α = .75; ωtotal = .79) in the correlated model expanded clinical sample (*N* = 1,200). With established suitability for factor analysis, CFA supported initial findings of a two-factor internal structure, demonstrating acceptable-to-good model fit (CFI = .96; TLI = .94; RMSEA = .07). Items 3 and 4 loaded onto the “school” factor, all other items load onto the “life” factor, mirroring findings from the normative sample and the original clinical sample. Notably, the item probing friend-related satisfaction had the weakest loading of .38 onto the “life” factor, as was reported in the normative sample. The correlated “life” and “school” structure were replicated in the clinical sample, although the weak loading of friend-related satisfaction and the two-item “school” factor warrant continued consideration.

### Measurement Invariance

In the first step to understand the NQLS’ ability to consistently measure the latent trait of QoL across samples, a baseline configural model was estimated across the normative and clinical samples using the correlated model. The unconstrained configural model, which specified the same correlated two-factor structure in both samples, demonstrated acceptable fit to the data across both samples χ*²* (26) = 170.68, CFI = .96; TLI = .94; RMSEA = .07 (90% CI [.06, .08]; SRMR = .04). Given the shared configural factor structure, we then evaluated for metric invariance by constraining the factor structure and item loadings. Evidence of metric invariance was found with ΔCFI ≤ .010, allowing us to test for scalar invariance.

Full scalar invariance with thresholds constrained was not supported by our data, showing that the NQLS may possess different thresholds for scoring in different settings (ΔCFI = .014). Items to be freed were identified through review of the standardized expected parameter change values. Freeing ‘NQLS_Life | t3’ brought the CFI change within the .010 limit (ΔCFI = .005) and allowed the partial scalar result to be used for subsequent comparisons. Evidence was not found to support partial strict invariance, though partial scalar invariance is sufficient for examination of latent means across samples.

Benjamini-Hochberg adjusted comparisons of latent means indicated that the clinical group scored significantly lower than the normative group on both the “life” (*M*diff = −.97, 90% CI [−1.06, −.88], *p* < .001) and the “school” factor (*M*diff = −.82, 90% CI [−.89, −.75], *p* < .001). We identified the normative group as the reference group and fixed its latent means and standardized variances; therefore, between-group differences were expressed as normative-group standard deviations (*M*diff). The clinical group scored .97 SD lower on the “life” factor and .82 SD lower on the “school” factor, indicating substantial differences in latent QoL across samples.

#### Normative and Clinical Sample Comparisons

Youth in the normative sample reported a mean NQLS total score of 19.96 (*SD* = 4.28), with item means ranging from 2.38 for school-related satisfaction to 3.18 for family-related satisfaction. In the clinical sample, all item level means were below 3.00, with a mean NQLS total score of 14.38 (*SD* = 4.74). School and grades showed the lowest reported satisfaction with means of 1.33 and 1.81, respectively, and family-related satisfaction received the highest average rating at 2.75.

Visualizing NQLS total scores in the clinical sample shows a nearly normal distribution, with slight item-level negative skew for satisfaction related to friends (γ = −.68) and family (γ = −.75), and slight positive skew for school (γ = .36). All other values fell within ± .15 of 0, with minimal kurtosis. In contrast, the normative sample demonstrated a negative skew and minimal kurtosis across all items as well as total score, consistent with the ceiling effect observed with high QoL in the normative sample. Figure 5 presents the differential distribution of NQLS total scores across both samples.

**Figure 5.**
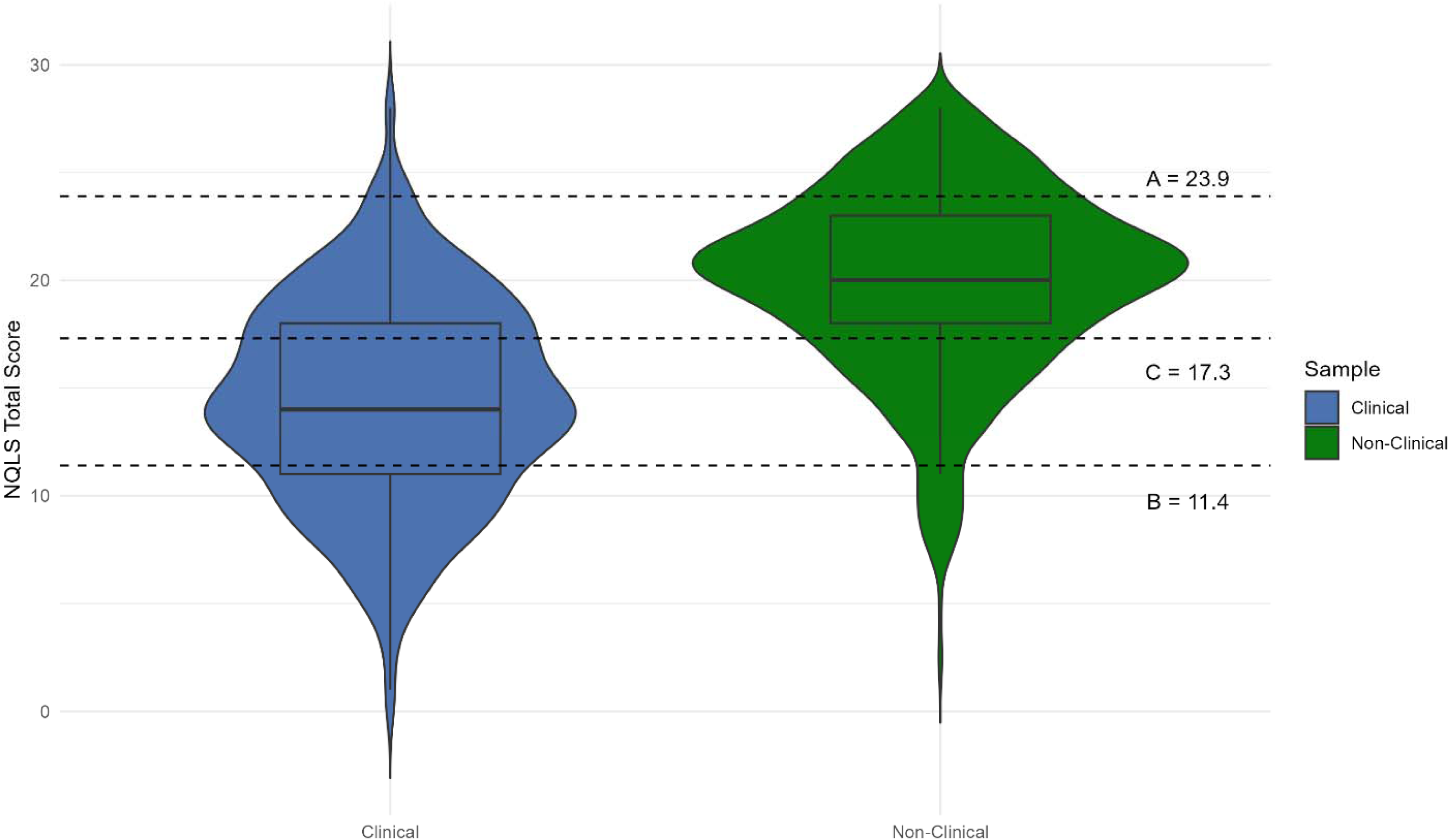
Nationwide Quality of Life Scale (NQLS) total score distribution by sample with clinical significance benchmarks.

Youth in the clinical sample reported significantly lower NQLS total scores than those in the normative sample (*t* = −29.05, *p* < .001, *d* = −1.23). The mean score for normative sample youth was more than 5 points higher than those in the clinical sample, placing the average clinical sample respondent in the 10^th^ percentile of the normative sample across all ages. This group-level comparison indicates the average clinical sample score was comparable to scores within the bottom 10% of the normative sample.

Across all QoL domains, normative sample means were significantly higher than those reported in the clinical sample after Benjamini-Hochberg correction (all *p*FDR < .001; Table 4). Between-sample differences ranged in size and magnitude. Friend-related satisfaction showed the lowest magnitude of difference between samples (*d* = .26), compared to health and life in general which showed the largest magnitude (both *d* = 1.10). Life showed the largest raw-score difference between samples (1.08 points), whereas friends showed the smallest difference (.27 points). These findings demonstrated substantial differentiation between normative and clinical-sample youth, particularly related to overall life, health, and school-related satisfaction.

**Table 4.** Average Nationwide Quality of Life Scale (NQLS) item scores by sample.

| **Quality of Life Domain** | **Normative Mean (SD)** | **Clinical Mean (SD)** | **Difference (Norm-Clin)** | **t-Score (df)** | **Effect Size (d)** |
| --- | --- | --- | --- | --- | --- |
| Friends | 2.94 (.90) | 2.67 (1.07) | 0.27 | 6.28 (2207) *** | 0.26 |
| Family | 3.18 (.79) | 2.75 (.98) | 0.43 | 11.50 (2204) *** | 0.48 |
| Grades | 2.80 (.98) | 1.81 (1.16) | 0.99 | 21.86 (2207) *** | 0.92 |
| School (in General) | 2.38 (1.09) | 1.33 (1.13) | 1.05 | 22.33 (2165) *** | 0.95 |
| Health | 2.99 (.84) | 2.00 (1.00) | 0.99 | 25.30 (2207) *** | 1.10 |
| Life | 2.89 (.82) | 1.81 (1.05) | 1.08 | 27.07 (2194) *** | 1.10 |
| Anticipated Future | 2.77 (.83) | 2.01 (1.05) | 0.76 | 19.06 (2198) *** | 0.80 |

*Note.* \*\*\* $p < .0005$ , two-tailed, after FDR correction. Positive differences indicate that youths in the normative sample reported higher QOL scores on average.

### Clinically Significant Change

National norms established in this study provide a reference for interpreting individual NQLS scores. For males, a total score of 20 would place them within the 50^th^ percentile of QoL and for females a total score of 21 corresponds to the 50^th^ percentile. The 50^th^ percentile is observed to be stable across all ages, meaning that this score equates to the median score regardless of age (Table 2).

A distribution-based minimally important difference (MID) was estimated as half of one standard deviation in the normative total score. With a normative *SD* of 4.28, the MID was estimated to be 2.14 points. Given that the NQLS is scored in whole points, a change of at least 3 points would be required to achieve the MID. The MID, a more liberal benchmark, is insightful for tracking QoL changes over time and is a preliminary indicator for potentially meaningful change, but it is not as well-known in psychotherapy applications in comparison to reliable change scores (Table 5). Not only is the clinical/normative total score difference statistically significant, but with a difference approximately 2.6 times the established MID, the magnitude of the group differentiation emerges.

**Table 5.** Clinical significance benchmarks and reliable change scores.

|  |  |
| --- | --- |
| *Descriptives (Full)* |  |
| Baseline *N* | 1008 |
| Mean (*SD*) | 19.96 (4.28) |
| Potential Range | 0–28 (7 items × 0–4) |
| Observed Range | 2–28 |
| $\omega_{\text{total}}$ | .88 |
| *Individual Change Thresholds (Consistency-Based)* |  |
| MID<sup>1</sup> (half-*SD*) | 2.14 |
| SEM<sup>2</sup> (using $\omega_{\text{total}}$) | 1.48 |
| 90% RCI<sup>3</sup> | 3.45 |
| 95% RCI | 4.11 |
| *Descriptives (Retest)* |  |
| Retest *N* | 90 |
| Baseline *M* (*SD*) | 19.51 (4.39) |
| Retest *M* (*SD*) | 20.29 (4.18) |
| Difference (Baseline - Retest) | –0.78 |
| *r* dependability | .73 |
| *Individual Change Thresholds (Dependability-Based)* |  |
| ICC(A,1)<sup>4</sup> | .718 [.597, .806] |
| Bland-Altman proportional-bias slope | 0.06, *p* = .50 (ns) |
| Johnson-Neyman region (gated to observed range) | [14.9, 21.3] |
| 90% RCI | 5.19 |
| 95% RCI | 6.19 |
| *Clinical Significance Benchmarks* |  |
| Away from clinical range (clinical *M* + 2*SD*)* | 23.9 |
| Crossing Closer (weighted *M*) threshold | 17.3 |
| Back into non-clinical range significance threshold (*M*–2*SD*)* | 11.4 |
| Empirical 5th percentile of general population | 12 |

*Note.* \*High NQLS score = higher QoL, therefore the equations for clinical significance benchmarks observed here are reversed from most symptom severity measures. Minimally important difference (MID)<sup>1</sup>, Standard error of the mean (SEM)<sup>2</sup>, Reliable change index (RCI)<sup>3</sup>, Intraclass correlation (ICC)<sup>4</sup>.

RC scores were estimated using both the more common internal consistency and the more conceptually preferable retest dependability metrics of the sample. This produces a substantially higher, more conservative bar for reliable change than an internal-consistency-only estimate would suggest and is the more conservative threshold for judging whether an individual’s change over time reflects genuine clinical improvement or deterioration rather than ordinary measurement noise. Importantly, the slight drift in scores observed at retest (see Test-Retest Dependability, above) does not meet the dependability-based reliable-change thresholds of 5.2 points at 90% confidence and 6.2 points at 95% confidence. These thresholds provide increasingly more stringent markers for clinical change in NQLS score. A 4-point change in score would achieve the MID, indicating that there is enough of a difference in the youth’s NQLS score for them to notice improvement. However, this does not reach the RC thresholds. The RC thresholds calculated with internal consistency indicate that the difference is big enough that it’s likely to be due to underlying change; the benefit of the retest dependability metric is its consideration of other reasons scores might change over a similar time interval. Using the whole point scale of the NQLS and our dependability metrics, a score change of 6 points meets the 90% RC threshold and a change of 7 points meets the 95% threshold, both of which are two points higher than RC thresholds calculated using internal consistency (4, 5, respectively; Table 5).

Based on the normative sample and our observed clinical sample, three clinical significance benchmarks were calculated to assist with evaluation of NQLS scores over time. The A, or “away,” benchmark of 23.9 identifies scores that are outside the range of scores typically observed in the clinical sample. A total score of 24 is required to meet the A benchmark, which indicates reported levels of QoL more than two standard deviations above the clinical mean. Youth with extremely low QoL, such as those in the observed clinical sample, may benefit from the application of the B benchmark at 11.4, which identifies that a score 12 or higher falls within two standard deviations of the normative mean, and they are moving “back” into the range of what could be expected within a normative sample. A score of at least 18 would indicate QoL concerns meeting at the C benchmark of 17.3, or “crossing” the estimated boundary between clinical and normative distributions. Total scores of 18 or higher are statistically more likely to come from the normative distribution than clinical distribution (Table 5).

## Discussion

Findings from the nationally representative BENCHMARK study suggest the NQLS is a reliable and valid measure of QoL in youth aged 12-18. National norms established for the NQLS place the average clinical sample score within the 10^th^ percentile of QoL reported by youth in the normative sample (μ = 14.4). We observe that youth from the normative sample have a total score 5.6 points higher than those of the clinical sample, and score significantly higher across all items compared to clinical-sample youth. This is consistent with the literature demonstrating decreased QoL in youth seeking treatment for MEB concerns (Celebre et al., 2021; Wever et al., 2024). The NQLS is well positioned for uptake in MBC systems and behavioral health research given its brevity, cross-sample reliability and validity, accessibility, and the clinical applications (norms, clinical significance benchmarks, reliable change thresholds) established in this study.

A score of 20 for males and 21 for females would place a respondent around the normative average, at the 50^th^ percentile. The stability of percentiles across age groups within the normative sample highlights the importance of intervention when low QoL is reported. The cross-sectional design of this study limits our ability to comment on the association of changes in age with natural changes in QoL, however, previous research has established the stability of QoL domains over time (Gillison et al., 2008; González-Blanch et al., 2018). Each percentile shows minimal change across age groups, with slight narrowing at the extremes (5^th^ and 95^th^ percentile). While females score higher than males across all timepoints, caution should be used interpreting a one-point difference on a scale with a total score range from 0-28.

We replicate the two-factor structural solution observed by Covarrubias and Fristad (2025) in the nationally representative sample, but offer additional guidelines for the interpretation of NQLS scores based on findings from the BENCHMARK sample. A two-factor correlated model and one-factor composite model were compared using EFA, CFA, GRM, and reliability estimates. Overall, fit indices are favorable for the correlated model, and evidence supports a two-factor internal structure of the NQLS containing “life” and “school” factors.

However, with only two items loading strongly onto the “school” factor many psychometric tests could not be performed; therefore, evidence for using the “school” factor as a standalone score is insufficient. Given the total score shows reliability, validity, precision, and stability, we recommend its use for clinical interpretations of NQLS scores. In research situations or large cross-group analyses, the two-factor correlated model should be used; however, the total score shows strong measurement properties and should be utilized when discussing an individual’s NQLS score or making clinical decisions.

No significant DIF was detected across sex, age, or race (White vs. non-White), which may support the presence of unbiased underlying trait differences, but sweeping generalizations should not be made about “typical” QoL between sexes, age groups, or racial/ethnic groups due to the restricted range of total score and, for race specifically, the modest non-White cell size (*n* = 172) relative to the sex and age comparisons. A finer-grained race/ethnicity analysis, such as separating specific groups rather than a White/non-White binary, is a natural extension for future work with a larger or differently stratified sample.

Factor loadings and fit indices for the “school” factor were stronger in the clinical sample compared to the normative sample. One possibility is that school-related satisfaction shows stronger differentiation among youth experiencing MEB problems, potentially in association with heightened school avoidance or school-related stress, leading to a more pronounced second factor (Fremont, 2003; Ingul et al., 2019). To avoid misattribution, if additional information is needed regarding an individual’s school performance, a more specific performance measure such as the VADRS performance subscale or the Adolescent Academic Problems Checklist (Sibley et al., 2014) should be administered.

NQLS total scores are most strongly associated with performance and functional impairment, while correlation with symptom severity measures show sufficient and expected convergent validity. The total score shows the widest range of conditional reliability, accurately identifying latent QoL with reliability above .80 for those between θ = −3.42 to θ = 1.57. The “life” factor shows its strongest association with internalizing symptoms and suicide-related constructs while the “school” factor is most strongly associated with performance. Both factor-level models show a narrower range of conditional reliability compared to the total score. Youth in the normative sample reported both higher NQLS raw scores and higher levels of the latent trait, QoL, compared to those in the clinical sample.

During treatment or progress monitoring, clinicians can utilize RC scores and MID to track within-person changes in youth QoL, using their own scores as progress markers. MID represents the amount of score change that a youth would likely notice, whereas RC denotes a large enough change that it is likely to reflect underlying improvement versus having a good day or week. RC thresholds may be particularly useful during the progress-monitoring phase of MBC, where scores are collected repeatedly to evaluate treatment response and inform care plans. Anchoring RC thresholds to the NQLS’s 2-week test-retest dependability correlation (*r* = .73) rather than to internal-consistency reliability (ωtotal = .88) allows us to capture genuine short-term fluctuation in reported QoL across occasions in a way that an internal-consistency estimate – computed from a single administration – structurally cannot (Revelle & Condon, 2019).

Individual scores can also be compared between-person against similar-aged peers using national norms and clinical significance benchmarks. When working with an individual, clinically significant improvement can be evaluated by considering both the magnitude of within-person change and the youth’s treatment status. For example, a seven-point increase in NQLS total score meets the threshold of reliable change with 95% confidence, indicating improvement beyond expected measurement error, while a total score of 18 crosses the C benchmark, indicating an individual’s movement from the clinical to normative side of the estimated distributional boundary. These benchmarks are interpretive aids for MBC implementation rather than diagnostic or treatment/eligibility cutoffs and should be integrated with symptom measures, functional outcomes, and information from the youth and family.

An inherent limitation of this study includes sampling biases from the administration of an online survey – average income and parental education is higher among respondents in the BENCHMARK sample than the national average and the sample likely unintentionally excludes families with little to no access to technology. Additionally, the sample contains fewer 18-year-olds than would be expected in a truly representative sample due to the nature of data collection. Youth and parents were polled in pairs to allow for dyadic comparisons, but 18-year-old respondents meet the age of majority and do not require a parent’s consent to answer surveys. This reduced the number of parents who reported on an 18-year-old, and therefore the number of 18-year-olds in the sample. The 18-year-old youth remained part of the sample as our study was sufficiently powered to detect results, but these youth are not analyzed independently of the full sample.

A notable strength offsetting these limitations is the study’s pre-registered 2-week retest panel (*N* = 90 after strict identity matching), which allowed reliable change thresholds to be anchored to a genuine test-retest dependability estimate – a design feature not available to most single-occasion normative studies, and one that yields a more conservative, defensible basis for individual-level change interpretation. Aside from the above departures, the normative sample is aligned with 2023 ACS estimates and represents the expected population of 12–18-year-old youth in the United States.

QoL measurement is part of a strengthening shift in the behavioral health field, where patient-centered outcomes are valued throughout treatment, and understood as integral to care planning and case conceptualization. Future research directions include understanding reporter differences in QoL and characteristics of disagreement, qualitative perspectives on the implementation and integration of QoL measurement from key stakeholders, further development of tools for the translation of research findings into clinical practice, and deeper analysis of the NQLS as it relates to other symptom severity measures. This study expands the research base and utility of the NQLS, provides interpretation guidelines for its scoring, clinical application, and measurement-based care integration, and poses future directions for researchers interested in QoL evaluation.

## CRediT roles

EAY conceptualized the project and secured provision of nationally representative data; YL and EN ran analyses and created visualizations; EAY cross-checked analyses and reporting using independent code; EN wrote all sections of the original draft; MAF provided supervision, resources, and clinical data; all authors reviewed and edited the full manuscript.

## Declaration of AI Use

Generative AI [ChatGPT Model 5.6 Sol] was used to 1) support code debugging in R, with all code checked and run by authors, Claude Code was used for an independent reanalysis and verification of the reproducibility of results; 2) copyedit the final manuscript draft; and 3) review adherence to standard BENCHMARK study procedures. An OpenEvidence search was used to look for any additional references pertaining to QoL measurement in youths and association with mental and behavioral health.

## Data availability statement

Code and sufficient data for replication available upon request from the corresponding author.

Disclosures

Dr. Eric A. Youngstrom is the co-founder and Executive Director of Helping Give Away Psychological Science, a 501c3; receives grant support from the National Institute of Health; he has received royalties from the American Psychological Association and Guilford Press, and he holds equity in Joe Startup Technologies. Dr. Mary A. Fristad receives grant support from the National Institute of Health, royalties from American Psychiatric Publishing and Guilford Press as well as editorial and travel support from the Society of Clinical Child and Adolescent Psychology. Ms. Elizabeth A. Nienaber and Ms. Yinuo Liu have no disclosures to report.

## Data Availability

Code and sufficient data for replication available upon request from the corresponding author.

## Acknowledgements

Supported by gifts from the Chlapaty, DiMarco, and Zipfel families to Nationwide Children’s Hospital to support the Institute for Mental and Behavioral Health Research (IMBHR). The authors gratefully acknowledge Jonathan Covarrubias for his assistance with the clinical dataset, and Marissa McClellan, Kevin Stephenson, Jacqui Pazaropoulos, Brittany Grate, and Kim Jones for their contributions to the development and implementation of the BENCHMARK project, including literature and reference review, study coordination and administration, data organization and management, data cleaning, and development of analytic resources.

