## Supplemental Materials for "Adolescent-Reported National Norms, Psychometrics, and Clinical Considerations for the Nationwide Quality of Life Scale (NQLS)"

**Author Note**

Elizabeth A. Nienaber <https://orcid.org/0009-0001-4585-4654>

Yinuo Liu <https://orcid.org/0009-0007-2606-575X>

Eric A. Youngstrom <https://orcid.org/0000-0003-2251-6860>

Mary A. Fristad <https://orcid.org/0000-0002-3189-2166>

Correspondence concerning this article should be addressed to Elizabeth A. Nienaber, Nationwide Children’s Hospital, 444 Butterfly Gardens Drive, Columbus, OH 43215, United States.

Table S1. Retest Sample Participant Demographics Table.

|  | Youth Retest (*N* = 90) |
| --- | --- |
| **Child Age** | 14.37 (1.67) |
| 12-13 years | 36.7% |
| 14-15 years | 33.3% |
| 16-18 years | 30.0% |
| **Child sex** | |
| Female | 51.1% |
| Male | 48.9% |
| **Race** | |
| White | 70.0% |
| Black or African American | 12.2% |
| Asian | 1.1% |
| American Indian or Alaska Native | 3.3% |
| Native Hawaiian or Pacific Islander | 1.1% |
| Some other race | 2.2% |
| Multiple races selected | 10.0% |
| **Ethnicity** | |
| Hispanic, Latino, or Spanish Origin | 17.1% |
| Not Hispanic, Latino, or Spanish Origin | 82.9% |
| **Parent/guardian education** | |
| High school graduate or higher | 100.0% |
| Bachelor’s degree or higher | 53.3% |
| **Median Household Income** | $70,000 - $79,999 |

**Figure S1.** Nationwide Gauge of Emotional Severity (N-Gauge) Measure.

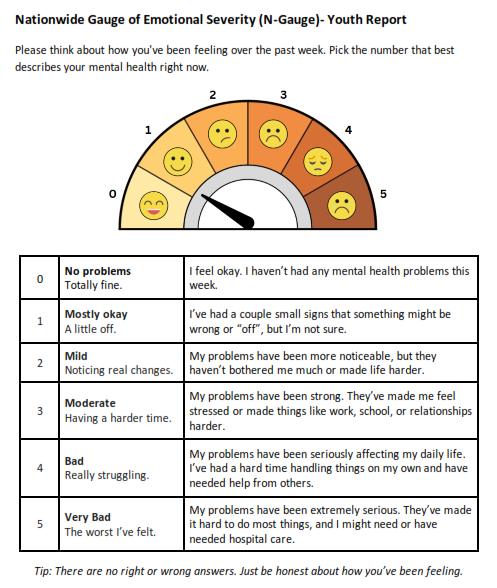

**Table S2.** GAMLSS model fit comparisons.

| **Model Distribution** | **Family Name** | ***df*** | **GAIC** | **BIC** | **Global Deviance** |
| --- | --- | --- | --- | --- | --- |
| Beta Binomial | BB | 1001 | 5743.68 | 5743.68 | 5709.11 |
| Box-Cox Cole Green | BCCG | 1000 | **5739.25** | **5739.25** | 5697.77 |
| Box-Cox Power Exponential | BCPE | 999 | 5742.65 | 5743.25 | **5695.25** |
| Normal | NO | 1001 | 5810.85 | 5810.85 | 5776.28 |

**Figure S2.** Box-Cox Cole Green (BCCG) worm plot.

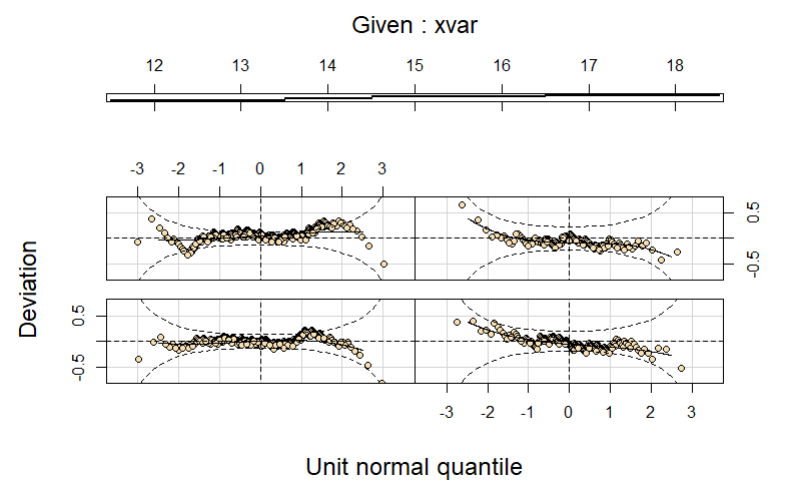

**Table S3.** Confirmatory Factor Analysis (CFA) and Exploratory Structural Equation Modeling (ESEM) Model Fit Overview.

|  | ***N*Factors** | ***χ^2^* (*df*)** | **CFI** | **TLI** | **RMSEA** | **SRMR** | **AIC*** | **BIC*** |
| --- | --- | --- | --- | --- | --- | --- | --- | --- |
| CFA | 1 | 414.26 (14) | .92 | .88 | .17 | .07 | 16490.62 | 16593.85 |
| CFA | 2 | 67.47 (13) | .99 | .98 | .07 | .03 | 16295.97 | 16404.11 |
| ESEM | 2 | 25.19 (8) | .99 | .99 | 005 | .02 | 16289.71 | 16422.44 |

*Note.* CFI and TLI scores closer to 1.0 are better. RMSEA and SRMR scores closer to zero indicate better fit. AIC and BIC are compared between models, and lower score wins. BIC differences >10 offer “strong evidence” in favor of that model (Raftery, 1995). ***AIC/BIC results from maximum likelihood estimation, not WLSMV.

**Table S4.** Factor loadings of the Nationwide Quality of Life Scale (NQLS) using exploratory factor analysis (EFA), confirmatory factor analysis (CFA), and exploratory structural equation modeling (ESEM).

|  | **EFA** | | | **CFA** | | | **ESEM** | |
| --- | --- | --- | --- | --- | --- | --- | --- | --- |
|  | **1F** | **2F** | | **1F - Composite** | **2F –**  **Correlated** | | **2F – Crossloading** | |
| **NQLS Item** | **‘QoL’** | **‘Life’** | **‘School’** | **‘QoL’** | **‘Life’** | **‘School’** | **‘Life’** | **‘School’** |
| Friends | 0.54 | 0.52 | - | 0.57 | 0.59 | - | 0.56 | 0.04 |
| Family | 0.62 | 0.66 | - | 0.70 | 0.72 | - | 0.71 | 0.01 |
| Grades | 0.55 | - | 0.77 | 0.63 | - | 0.74 | -0.03 | 0.80 |
| School in General | 0.62 | - | 0.73 | 0.69 | - | 0.86 | 0.09 | 0.76 |
| Health | 0.56 | 0.55 | - | 0.62 | 0.64 | - | 0.59 | 0.05 |
| Life | 0.82 | 0.93 | - | 0.90 | 0.93 | - | 0.96 | -0.05 |
| (Anticipated) Future | 0.64 | 0.55 | - | 0.69 | 0.70 | - | 0.60 | 0.13 |

***Note.*** All analyses used polychoric correlation matrices. EFA used promax rotation.

**Figure S3.** Measurement model diagram and loadings for the exploratory structural equation model (ESEM).

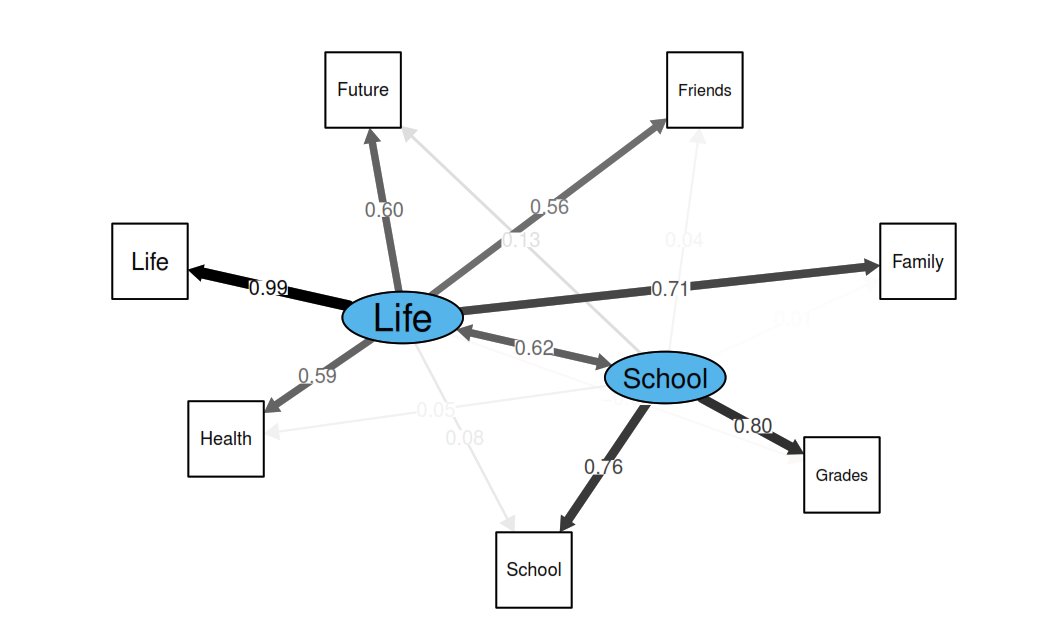

**Table S5.** Comparison of NQLS factor-level correlation with symptom severity measures.

| **Measure** | ***r_1_*** | ***r_2_*** | **Cohen's *q*** | **Effect Size** | **Steiger's *Z*** | ***p*FDR** | **Significance** |
| --- | --- | --- | --- | --- | --- | --- | --- |
| VQIDS Depression | -0.53 | -0.37 | -0.20 | Small | -5.77 | <.001 | *** |
| CHRT Suicide Risk +AF^1^ | -0.55 | -0.40 | -0.19 | Small | -5.64 | <.001 | *** |
| CHRT Propensity | -0.41 | -0.36 | -0.06 | - | -1.95 | 0.069 | - |
| CHRT Suicidal Thoughts | -0.55 | -0.39 | -0.20 | Small | -6.01 | <.001 | *** |
| PSC-17 General MEB^2^ Problems | -0.51 | -0.39 | -0.14 | Small | -4.20 | <.001 | *** |
| PSC-17 Internalizing | -0.53 | -0.37 | -0.20 | Small | -5.88 | <.001 | *** |
| PSC-17 Externalizing | -0.38 | -0.32 | -0.07 | - | -2.06 | 0.057 | - |
| PSC-17 Attention | -0.36 | -0.30 | -0.08 | Small | -2.36 | 0.030 | * |
| VADRS ADHD | -0.41 | -0.34 | -0.07 | Small | -2.24 | 0.039 | * |
| VADRS Hyperactivity/Impulsivity | -0.30 | -0.21 | -0.10 | Small | -3.06 | 0.004 | ** |
| VADRS Inattention | -0.44 | -0.41 | -0.04 | - | -1.11 | 0.318 | - |
| VADRS Performance | -0.54 | -0.63 | 0.14 | Small | 3.78 | <.001 | *** |
| N-Gauge Emotional Severity | -0.44 | -0.34 | -0.12 | Small | -3.66 | <.001 | *** |

*Note.* ^1^Suicide Risk and Associated Factors. ^2^Mental, emotional, and behavioral (MEB). Cohen’s *q* was calculated as the difference between Fisher-transformed correlations, with the sign corresponding to the order of scores listed in the comparison. Absolute values of .10, .30, and .50 conventionally represent small, medium, and large differences, respectively. Steiger’s test was used to compare dependent correlations sharing a common variable. Reported *p* values were adjusted for multiple comparisons using the Benjamini–Hochberg false discovery rate procedure. *p*<.05, ***p* <.005, ****p* <.0005, after FDR correction.

**Table S6.** Graded response model fit indices.

| **Model** | **C_2_ (df)** | ***M_2_*** | **CFI** | **TLI** | **RMSEA [90% CI]** | **SRMSR** | **AIC** | **BIC** | **SABIC** |
| --- | --- | --- | --- | --- | --- | --- | --- | --- | --- |
| Correlated | 133.25 (13) | 48.83 | .99 | .98 | .06 [.06-.07] | .03 | 15041.78 | 15218.75 | 15104.41 |
| Composite | 550.82 (14) | 224.89 | .94 | .90 | .12 [.12-.14] | .07 | 15226.65 | 15397.70 | 15286.54 |

**Table S7.** Item-level fit statistics for 2-factor correlated GRM model

| **Item** | ***a_1_*** | ***a_2_*** | ***b_1_*** | ***b_2_*** | ***b_3_*** | ***b_4_*** | **RMSD** |
| --- | --- | --- | --- | --- | --- | --- | --- |
| Friends | 0.92 | 0 | -4.15 | -2.45 | -1.07 | 1.48 | 0.05 |
| Family | 1.41 | 0 | -3.34 | -2.27 | -1.07 | 0.89 | 0.06 |
| Grades | 0 | 2.20 | -1.67 | -0.77 | 0.06 | 1.36 | 0.08 |
| School | 0 | 3.38 | -0.95 | -0.31 | 0.44 | 1.71 | 0.11 |
| Health | 1.98 | 0 | -2.26 | -1.16 | -0.15 | 1.39 | 0.05 |
| Life | 4.32 | 0 | -1.52 | -0.84 | -0.01 | 1.33 | 0.09 |
| Future | 1.96 | 0 | -2.11 | -1.27 | 0.15 | 1.53 | 0.06 |
